# Accuracy of Rapid Antigen Testing across SARS-CoV-2 Variants

**DOI:** 10.1101/2022.03.21.22272279

**Authors:** Paul K. Drain, Meagan Bemer, Jennifer F. Morton, Ronit Dalmat, Hussein Abdille, Katherine Thomas, Timsy Uppal, Derrick Hau, Heather R. Green, Marcellene A. Gates-Hollingworth, David P. AuCoin, Subhash C. Verma

**Affiliations:** Departments of Global Health, Medicine, and Epidemiology, University of Washington, Seattle; Department of Microbiology and Immunology, Reno School of Medicine, University of Nevada, Reno

## Abstract

Variants of SARS-CoV-2 have mutations in the viral genome that may alter the accuracy of rapid diagnostic tests. We conducted analytical and clinical accuracy studies of two FDA-approved rapid antigen tests—SCoV-2 Ag *Detect*™ Rapid Test (InBios International, Seattle) and BinaxNOW™ COVID-19 Ag CARD; (Abbott Laboratories, Chicago)—using three using replication-competent variants or strains, including Omicron (B.1.1.529/BA.1), Delta (B.1.617.2), and a wild-type of SARS-CoV-2 (USA-WA1/2020). Overall, we found non-significant differences in the analytical limit of detection or clinical diagnostic accuracy of rapid antigen testing across SARS-CoV-2 variants. This study provides analytical and clinical performance data to demonstrate the preserved accuracy of rapid antigen testing across SARS-CoV-2 variants among symptomatic adults.

---

Variants of SARS-CoV-2 have mutations in the viral genome that may alter the accuracy of rapid diagnostic tests.^1^ Molecular tests can be affected by single point mutations, whereas antigen tests may require multiple mutations to change the confirmation of viral protein epitopes. The Omicron variant has numerous mutations in the spike and nucleocapsid proteins,^2^ which has raised concerns about the analytical and clinical accuracy of rapid antigen testing.^3^

We conducted an analytical accuracy study of two FDA-approved rapid antigen tests—SCoV-2 Ag *Detect*™ Rapid Test (InBios International, Seattle) and BinaxNOW™ COVID-19 Ag CARD; (Abbott Laboratories, Chicago)—using three using replication-competent variants or strains, including Omicron (B.1.1.529/BA.1), Delta (B.1.617.2), and a wild-type of SARS-CoV-2 (USA-WA1/2020).

Omicron SARS-CoV-2 virus was isolated from a nasal swab collected in Maryland on November 27, 2021 and obtained through BEI Resources. The virus was cultured on *Cercopithecus aethiops* kidney cells (VERO E6) expressing transmembrane protease serine 2 gene (TMPRSS2) and angiotensin-converting enzyme 2 (ACE2), VERO E6-ACE2 and TMPRSS2. Variant identification of the cultured virus was confirmed through whole genome sequencing and cross-referenced with GISAID (Global Initiative on Sharing Avian Influenza Data, accession number, EPI_ISL_7160424). The replication competence of the virus stock was determined by measuring 50% tissue culture infectious dose (TCID_50_). A corresponding quantitative measure of infectious viral particles was determined by assaying the plaque forming units (PFUs), through infection of VERO E6-ACE2 and TMPRSS2, using a standard plaque assay protocol.^4^ Similarly, we used the Delta (B.1.617.2) variant (GISAID: EPI_ISL_2103264) and USA-WA1/2020 strain (GenBank: MN985325.1) of SARS-CoV-2, which have known TCID_50_ values of 1.1 ×10^6^ and 1.6 ×10^6^, respectively, for comparative analytical testing.

Performance and the analytical limit of detection for Omicron detection were measured by spiking negative clinical nasal swab matrices with replication-competent virus to establish three stock viral concentrations of 5.0 ×10^4^, 1.25 ×10^4^, and 3.12 ×10^3^ TCID_50_/mL. From stock concentrations, we transferred 20 µL of sample onto nasal swabs to generate high, medium, and low viral concentrations of 1,000, 250, 62.5 TCID_50_ per swab, respectively. We tested the viral dilutions across variants in triplicate for both the SCoV-2 Ag *Detect*™ Rapid Test (Figure S1) and BinaxNOW™ COVID-19 Ag CARD (Figure S2). Based on the visual signal intensity, the limit of detection for these rapid nucleocapsid antigen tests was approximately 62.5 TCID_50_, which is equivalent to 51 PFUs (Figure S3).

To complement the analytical accuracy study of the SCoV-2 Ag *Detect*™ Rapid Test, we further conducted a clinical diagnostic accuracy study among 802 participants at multiple testing locations in King County, Washington from February 2021—January 2022, during three distinct phases of SARS-CoV-2 infections (pre-Delta, Delta, Omicron) (Table S1). Participants were ≥18 years old reporting onset of Covid-19-like symptoms within the prior five days. We collected two anterior nasal swabs—one for on-site testing by the SCoV-2 Ag *Detect*™ Rapid Test and one for reverse transcriptase polymerase chain reaction (rt-PCR) testing. The study received ethical approval from the University of Washington (STUDY00009981), and participants provided verbal informed consent.

Overall, we found non-significant differences in the analytical limit of detection or clinical diagnostic accuracy of rapid antigen testing across SARS-CoV-2 variants (Table 1). The positive percent agreement ranged from 81-91%, with improved sensitivity for swabs with a lower cycle threshold, which correlates to a higher viral load (Table S2). Visual signals were positively associated with viral concentration and had more variation at low concentrations (Figure S1 and S2). We found no real differences for clinical test performance by vaccination status or days since symptom onset (Table S2). The negative percent agreement was high across study periods.

**Table 1.** Analytical and clinical accuracy of SCoV-2 Ag *Detect*™ Rapid Test across SARS-CoV-2 variants.

| Predominant Variant | Pre-Delta<sup>1</sup> | Delta | Omicron | Total<sup>2</sup> |
| --- | --- | --- | --- | --- |
| Testing Period<sup>3</sup> | Feb 17–April 29, 2021 | Sept 2–Nov 30, 2021 | Dec 13, 2021–Jan 11, 2022 | Feb 2021 – Jan 2022 |
| Covid-19 positive<sup>4</sup> / Participants with valid rt-PCR results | 64 / 296 | 43 / 289 | 73 / 212 | 180 / 797 |
| Days since symptom onset, mean ±SD | 2.2 ±1.2 | 2.3 ±1.2 | 2.5 ±1.3 | 2.3 ±1.2 |
| Cycle threshold values among SARS-CoV-2 positive by rt-PCR, mean ±SD | 23.9 ±5.2 | 27.6 ±4.6 | 28.0 ±4.8 | 26.5 ±5.3 |
| Positive percent agreement for rapid antigen test, % (95% CI)<sup>5</sup> | 81.2 (69.5, 89.9) | 90.7 (77.9, 97.4) | 83.6 (73.0, 91.2) | 84.4 (78.3, 89.4) |
| Negative percent agreement for rapid antigen test, % (95% CI)<sup>5</sup> | 100 (98.4, 100) | 99.6 (97.8, 100) | 100 (97.4, 100) | 99.8 (99.1, 100) |
| Analytical limit of detection for rapid antigen test (TCID<sub>50</sub> per swab) | 62.5 | 62.5 | 62.5 | 62.5 |

CI=confidence interval; SD=standard deviation
1. The Pre-Delta phase consisted mostly of Alpha, Epsilon, and Gamma variants in this study population 2. Five participants (3 from Delta phase, 2 from Omicron phase) were missing rt-PCR results and excluded from analyses. 3. Testing periods were determined based on Washington State's Department of Health data on the circulation of predominant variants (https://www.doh.wa.gov/Portals/1/Documents/1600/coronavirus/data-tables/420-316-SequencingAndVariantsReport.pdf). 4. Covid-19 positive was defined as a person who reported Covid-19-like symptoms and had a positive lab-based rt-PCR test for SARS-CoV-2. 5. 95% CIs computed using the exact binomial (Clopper-Pearson) method.

This study provides analytical and clinical performance data to demonstrate the preserved accuracy of rapid antigen testing across SARS-CoV-2 variants among symptomatic adults. Other studies have similarly demonstrated good analytical sensitivity for rapid antigen tests to detect Omicron,^5,6^ and with similar limits of analytical detection between the Omicron variant and USA-WA1/2020 strain.^7^ A field study has shown the BinaxNOW™ COVID-19 Ag CARD to have good clinical accuracy for Omicron.^8^ Our study has demonstrated both the analytical and clinical accuracy of rapid antigen testing across several variants of concern, including Omicron and Delta.

A study strength was detecting circulating variants representative of the community prevalence over a 12-month period, and additional clinical and analytical comparative studies may still be warranted for other rapid antigen tests. Since rapid antigen tests may correlate with recovery of replication-competent SARS-CoV-2^9^ and appear to retain accuracy across variants, ongoing home-based rapid antigen testing programs may be an important intervention to reduce global SARS-CoV-2 transmission.

## Data Availability

All data produced in the present work are contained in the manuscript

## Funding and Acknowledgements

The study was supported by InBios International Inc., which had no role in the data analyses, interpretation, or reporting of these results. Dr. Drain reports receiving grant support, paid to his institution, from the National Institutes of Health, the Centers for Disease Control and Prevention, and the Bill and Melinda Gates Foundation. No other potential conflict of interest relevant to this article was reported.

SARS-CoV-2 isolate USA-WA1/2020 was deposited by the Centers for Disease Control and Prevention and obtained through BEI Resources, NIAID, NIH: SARS-Related Coronavirus 2, NR-52281. The following reagents were obtained through BEI Resources, NIAID, NIH: SARS-Related Coronavirus 2, Isolate hCoV-19/USA/MD-HP05285/2021 (Lineage B.1.617.2; Delta Variant), NR-55671, Isolate hCoV-19/USA/MD-HP20874/2021 (Lineage B.1.1.529; Omicron Variant), NR-56461, contributed by Dr. Andrew Pekosz.

---

**Supplemental Table 1.** Characteristics of participants enrolled in the clinical accuracy study.

|  | **Pre-Delta** | **Delta** | **Omicron** | **All participants** |
| --- | --- | --- | --- | --- |
|  | **N (%)** | **N (%)** | **N (%)** | **N (%)** |
| Total | 296 | 292 | 214 | 802 |
| Sex at birth<sup>1</sup> |  |  |  |  |
| Female | 160 (54.1) | 191 (65.4) | 116 (54.2) | 467 (58.2) |
| Male | 135 (45.6) | 101 (34.6) | 98 (45.8) | 334 (41.6) |
| Gender<sup>1</sup> |  |  |  |  |
| Woman | 160 (54.1) | 184 (63.0) | 118 (55.1) | 462 (57.6) |
| Man | 134 (45.3) | 103 (35.3) | 94 (43.9) | 331 (41.3) |
| Nonbinary or genderqueer | 1 (0.3) | 5 (1.7) | 1 (0.5) | 7 (0.9) |
| Prefer not to answer | 0 (0) | 0 (0) | 1 (0.5) | 1 (0.1) |
| Race and Ethnicity<sup>1</sup> |  |  |  |  |
| American Indian or Alaskan Native | 6 (2.0) | 3 (1.0) | 1 (0.5) | 10 (1.2) |
| Native Hawaiian or Pacific Islander | 9 (3.0) | 12 (4.1) | 8 (3.7) | 29 (3.6) |
| Hispanic or Latinx | 52 (17.6) | 33 (11.3) | 27 (12.6) | 112 (14.0) |
| Asian | 27 (9.1) | 18 (6.2) | 20 (9.3) | 65 (8.1) |
| White | 149 (50.3) | 167 (57.2) | 116 (54.2) | 432 (53.9) |
| Black or African American | 27 (9.1) | 37 (12.7) | 23 (10.7) | 87 (10.8) |
| More than one race | 12 (4.1) | 19 (6.5) | 12 (5.6) | 43 (5.4) |
| A race or ethnicity not listed | 6 (2.0) | 1 (0.3) | 1 (0.5) | 8 (1.0) |
| Prefer not to answer | 7 (2.4) | 2 (0.7) | 6 (2.8) | 15 (1.9) |
| Age in years (mean ±SD) | 34.3 ±11.0 | 38.4 ±13.5 | 39.8 ±15.0 | 37.3 ±13.3 |
| Contact with COVID+ person within prior 2 weeks |  |  |  |  |
| No | 182 (61.5) | 190 (65.1) | 105 (49.1) | 477 (59.5) |
| Yes | 75 (25.3) | 75 (25.7) | 89 (41.6) | 239 (29.8) |
| Unknown | 39 (13.2) | 27 (9.2) | 20 (9.3) | 86 (10.7) |
| Symptoms |  |  |  |  |
| Days since symptom onset (median [IQR]) | 2 [2] | 2 [2] | 2 [1] | 2 [2] |
| COVID-19 Vaccination<sup>2</sup> |  |  |  |  |
| Not vaccinated | 296 (100) | 95 (32.5) | 33 (15.4) | 424 (52.9) |
| One dose | 0 (0) | 24 (8.2) | 12 (5.6) | 36 (4.5) |
| Two doses | 0 (0) | 165 (56.5) | 110 (51.4) | 275 (34.3) |
| Three doses | 0 (0) | 7 (2.4) | 59 (27.6) | 66 (8.2) |
| Swab collection for all rt-PCR results |  |  |  |  |
| Supervised self-collection | 296 (100) | 41 (14.0) | 214 (100) | 551 (68.7) |
| Collected by research coordinator | 0 (0) | 251 (86.0) | 0 (0) | 251 (31.3) |
| Swab collection among rt-PCR positives |  |  |  |  |
| Supervised self-collection | 64 (100) | 5 (11.6) | 73 (100) | 142 (78.9) |
| Collected by research coordinator | 0 (0) | 38 (88.4) | 0 (0) | 38 (21.1) |
| Cycle threshold values by rt-PCR among SARS-CoV-2 positive (median [IQR]) | 22.5 [5.9] | 27.1 [5.6] | 27.1 [5.6] | 26.2 [6.6] |

IQR=interquartile range; rt-PCR=reverse transcriptase polymerase chain reaction; SD=standard deviation.
1. Sex assigned at birth, gender, and race/ethnicity are not available for one pre-delta participant. Woman included responses "woman" and "transgender woman"; man included responses "man" and "transgender man".
2. Vaccination status was not available for one Delta participant.

**Supplemental Table 2.** Clinical accuracy of SCoV-2 Ag *Detect*™ Rapid Test across SARS-CoV-2 variants, as stratified by days since symptom onset, vaccination status, and cycle threshold values by rt-PCR.

|  | N<sup>1</sup> | TP/(TP+FN) | PPA (95% CI)<sup>2</sup> | TN/(TN+FP) | NPA (95% CI)<sup>2</sup> |
| --- | --- | --- | --- | --- | --- |
| **All participants** | **797** | **152/180** | **84.4 (78.3, 89.4)** | **616/617** | **99.8 (99.1, 100)** |
| Days since symptom onset |  |  |  |  |  |
| 0-3 days | 662 | 119/141 | 84.4 (77.3, 90.0) | 520/521 | 99.8 (98.9, 100) |
| 4-5 days | 135 | 33/39 | 84.6 (69.5, 94.1) | 96/96 | 100 (96.2, 100) |
| Vaccination status |  |  |  |  |  |
| No doses of vaccine | 423 | 77/91 | 84.6 (75.5, 91.3) | 332/332 | 100 (98.9, 100) |
| At least one dose of vaccine | 373 | 75/89 | 84.3 (75.0, 91.1) | 283/284 | 99.6 (98.1, 100) |
| N gene Ct value by rt-PCR |  |  |  |  |  |
| Ct ≤ 33 | 155 | 146/155 | 94.2 (89.3, 97.3) | 0/0 | NC |
| Ct > 33 | 642 | 6/25 | 24.0 (9.4, 45.1) | 616/617 | 99.8 (99.1, 100) |
| Ct ≤ 30 | 145 | 142/145 | 97.9 (94.1, 99.6) | 0/0 | NC |
| Ct > 30 | 652 | 10/35 | 28.6 (14.6, 46.3) | 616/617 | 99.8 (99.1, 100) |
| **Pre-Delta** | **296** | **52/64** | **81.2 (69.5, 89.9)** | **232/232** | **100 (98.4, 100)** |
| Days since symptom onset |  |  |  |  |  |
| 0-3 days | 254 | 44/53 | 83.0 (70.2, 91.9) | 201/201 | 100 (98.2, 100) |
| 4-5 days | 42 | 8/11 | 72.7 (39.0, 94.0) | 31/31 | 100 (88.8, 100) |
| Vaccination status |  |  |  |  |  |
| No doses of vaccine | 296 | 52/64 | 81.2 (69.5, 89.9) | 232/232 | 100 (98.4, 100) |
| At least one dose of vaccine | 0 | 0/0 | NC | 0/0 | NC |
| N gene Ct value by rt-PCR |  |  |  |  |  |
| Ct ≤ 33 | 58 | 51/58 | 87.9 (76.7, 95.0) | 0/0 | NC |
| Ct > 33 | 238 | 1/6 | 16.7 (0.4, 64.1) | 232/232 | 100 (98.4, 100) |
| Ct ≤ 30 | 54 | 51/54 | 94.4 (84.6, 98.8) | 0/0 | NC |
| Ct > 30 | 242 | 1/10 | 10.0 (0.3, 44.5) | 232/232 | 100 (98.4, 100) |
| **Delta** | **289** | **39/43** | **90.7 (77.9, 97.4)** | **245/246** | **99.6 (97.8, 100)** |
| Days since symptom onset |  |  |  |  |  |
| 0-3 days | 244 | 32/36 | 88.9 (73.9, 96.9) | 207/208 | 99.5 (97.4, 100) |
| 4-5 days | 45 | 7/7 | 100 (59.0, 100) | 38/38 | 100.0 (90.7, 100) |
| Vaccination status |  |  |  |  |  |
| No doses of vaccine | 94 | 12/12 | 100 (73.5, 100) | 82/82 | 100.0 (95.6, 100) |
| At least one dose of vaccine | 194 | 27/31 | 87.1 (70.2, 96.4) | 162/163 | 99.4 (96.6, 100) |
| N gene Ct value by rt-PCR |  |  |  |  |  |
| Ct ≤ 33 | 38 | 37/38 | 97.4 (86.2, 99.9) | 0/0 | NC |
| Ct > 33 | 251 | 2/5 | 40.0 (5.3, 85.3) | 245/246 | 99.6 (97.8, 100) |
| Ct ≤ 30 | 35 | 35/35 | 100 (90.0, 100) | 0/0 | NC |
| Ct > 30 | 254 | 4/8 | 50.0 (15.7, 84.3) | 245/246 | 99.6 (97.8, 100) |
| **Omicron** | **212** | **61/73** | **83.6 (73.0, 91.2)** | **139/139** | **100 (97.4, 100)** |
| Days since symptom onset |  |  |  |  |  |
| 0-3 days | 164 | 43/52 | 82.7 (69.7, 91.8) | 112/112 | 100 (96.8, 100) |
| 4-5 days | 48 | 8/21 | 85.7 (63.7, 97.0) | 27/27 | 100.0 (87.2, 100) |
| Vaccination status |  |  |  |  |  |
| No doses of vaccine | 33 | 13/15 | 86.7 (59.5, 98.3) | 18/18 | 100 (81.5, 100) |
| At least one dose of vaccine | 179 | 48/58 | 82.8 (70.6, 91.4) | 121/121 | 100 (97.0, 100) |
| N gene Ct value by rt-PCR |  |  |  |  |  |
| Ct ≤ 33 | 59 | 58/59 | 98.3 (90.9, 100) | 0/0 | NC |
| Ct > 33 | 153 | 3/14 | 21.4 (4.7, 50.8) | 139/139 | 100 (97.4, 100) |
| Ct ≤ 30 | 56 | 56/56 | 100 (93.6, 100) | 0/0 | NC |
| Ct > 30 | 156 | 5/17 | 29.4 (10.3, 56.0) | 139/139 | 100 (97.4, 100) |

Ct=cycle threshold; FN=false negative; FP=false positive; NC=not calculable; NPA=negative percent agreement; PPA=positive percent agreement; rt-PCR=reverse transcriptase polymerase chain reaction; TN=true negative; TP=true positive.
1. Five participants (3 from Delta phase, 2 from Omicron phase) were missing rt-PCR results and excluded from analyses.
2. 95% CIs computed using the exact binomial (Clopper-Pearson) method.

**Supplemental Figure 1.**
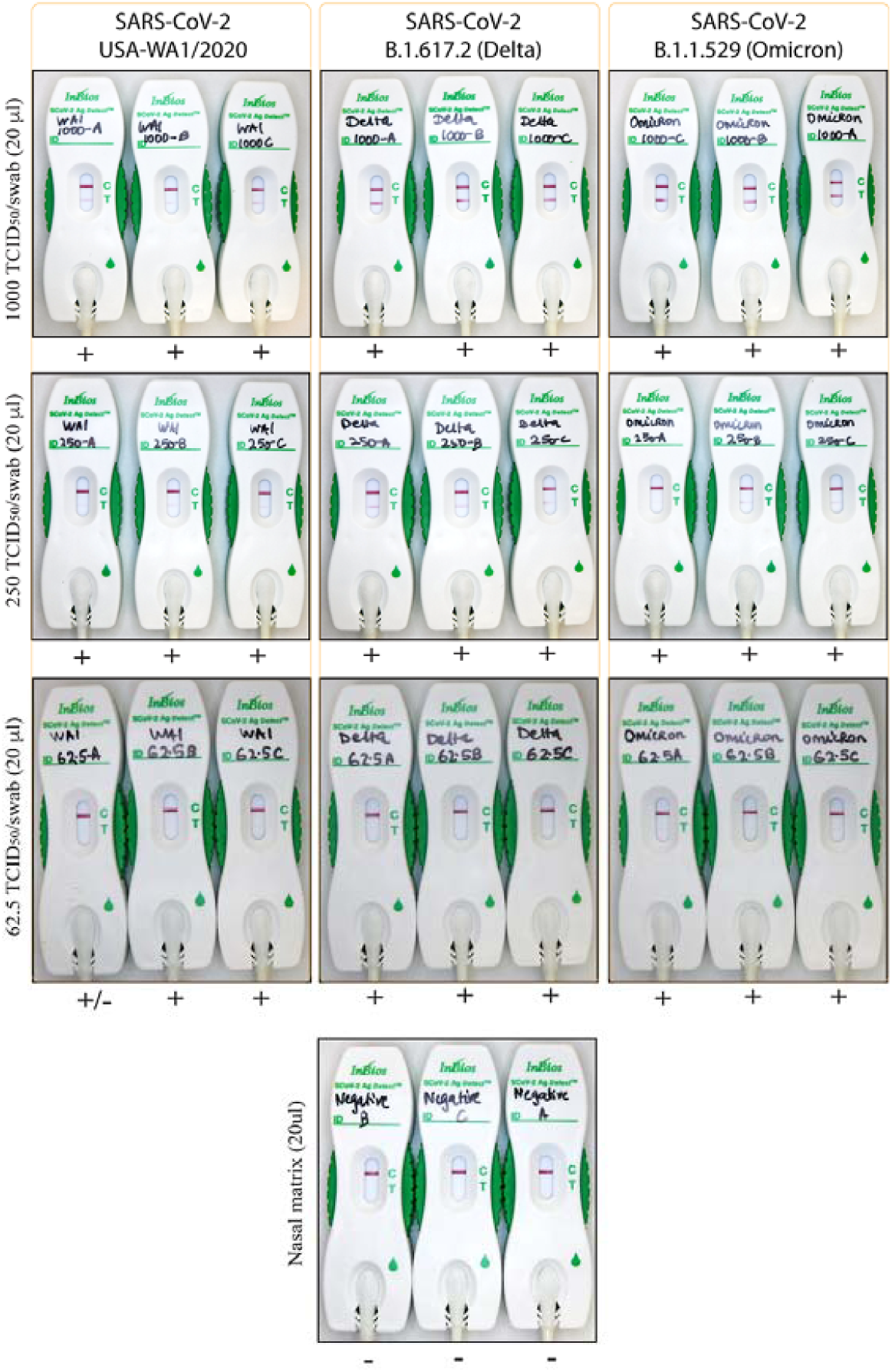
Images of the SCoV-2 Ag *Detect*™ Rapid Test across SARS-CoV-2 variants and stratified by viral load of TCID_50_ per swab. After establishing stock viral concentrations of 5.0 ×10^4^, 1.25 ×10^4^, and 3.12 ×10^3^ TCID_50_/mL, we transferred 20 µL of sample onto nasal swabs to generate high, medium, and low viral concentrations of 1,000, 250, 62.5 TCID_50_ per swab, respectively. We tested the viral dilutions across variants in triplicate using a SCoV-2 Ag *Detect*™ Rapid Test to establish the analytical limit of detection.

**Supplemental Figure 2.**
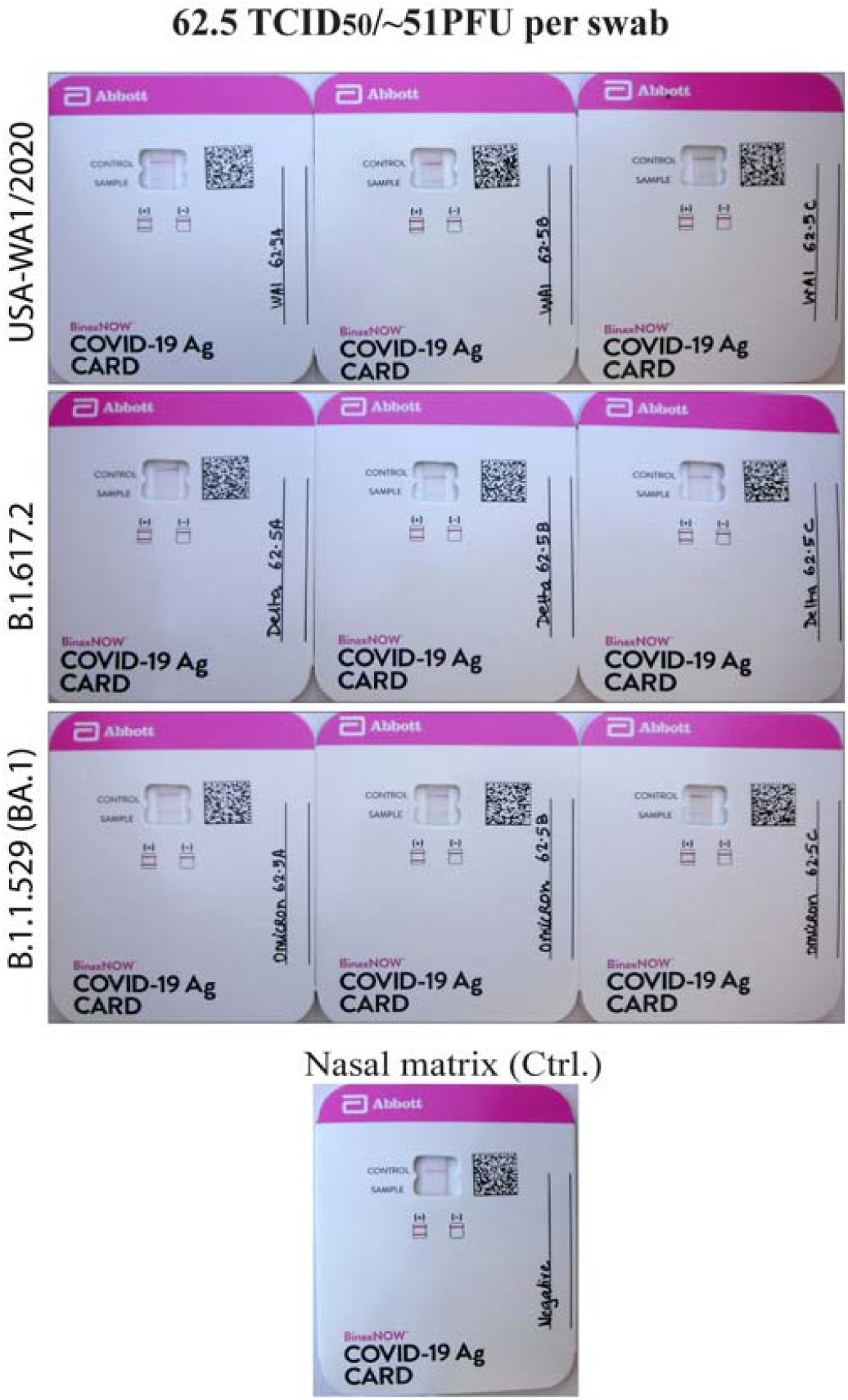
Images of the BinaxNOW COVID-19 Ag CARD across SARS-CoV-2 variants at 62.5 TCID_50_ viral load per swab.

**Supplemental Figure 3.**
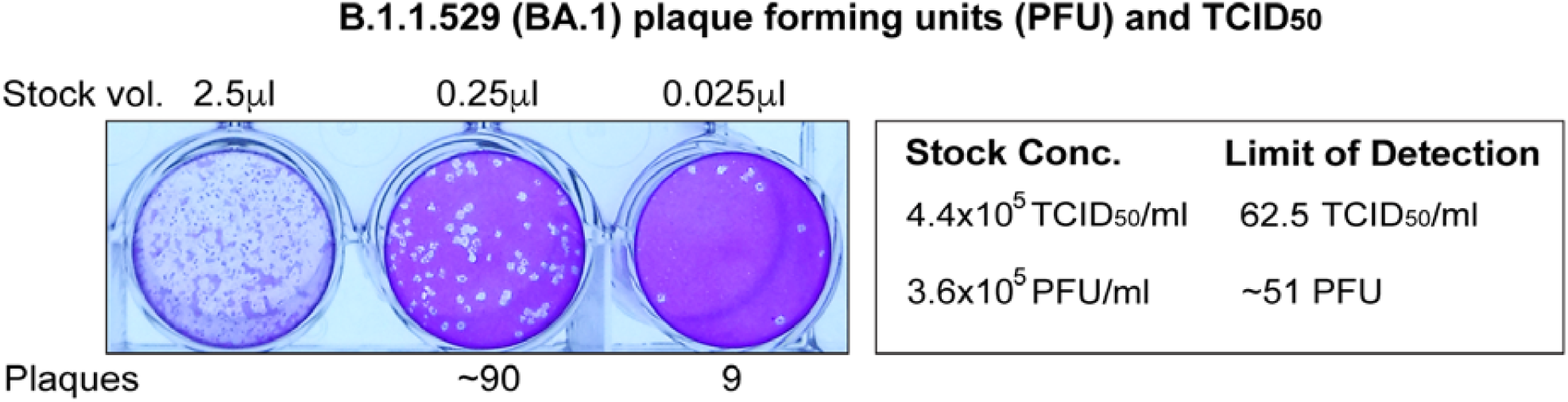
Plaque Assay to determine the replication competence of the SARS-CoV-2 isolate hCoV-19/USA/MD-HP20874/2021 (Lineage B.1.1.529; Omicron Variant).

